# Virological and serological kinetics of SARS-CoV-2 Delta variant vaccine-breakthrough infections: a multi-center cohort study

**DOI:** 10.1101/2021.07.28.21261295

**Authors:** Po Ying Chia, Sean Wei Xiang Ong, Calvin J Chiew, Li Wei Ang, Jean-Marc Chavatte, Tze-Minn Mak, Lin Cui, Shirin Kalimuddin, Wan Ni Chia, Chee Wah Tan, Louis Yi Ann Chai, Seow Yen Tan, Shuwei Zheng, Raymond Tzer Pin Lin, Linfa Wang, Yee-Sin Leo, Vernon J Lee, David Chien Lye, Barnaby Edward Young

## Abstract

**Objectives:** Highly effective vaccines against severe acute respiratory syndrome coronavirus 2 (SARS-CoV-2) have been developed but variants of concerns (VOCs) with mutations in the spike protein are worrisome, especially B.1.617.2 (Delta) which has rapidly spread across the world. We aim to study if vaccination alters virological and serological kinetics in breakthrough infections.

**Methods:** We conducted a multi-centre retrospective cohort study of patients in Singapore who had received a licensed mRNA vaccine and been admitted to hospital with B.1.617.2 SARS-CoV-2 infection. We compared the clinical features, virological and serological kinetics (anti-nucleocapsid, anti-spike and surrogate virus neutralization titres) between fully vaccinated and unvaccinated individuals.

**Results:** Of 218 individuals with B.1.617.2 infection, 84 had received a mRNA vaccine of which 71 were fully vaccinated, 130 were unvaccinated and 4 received a non-mRNA. Despite significantly older age in the vaccine breakthrough group, the odds of severe COVID-19 requiring oxygen supplementation was significantly lower following vaccination (adjusted odds ratio 0.07 95%CI: 0.015-0.335, p=0.001). PCR cycle threshold (Ct) values were similar between both vaccinated and unvaccinated groups at diagnosis, but viral loads decreased faster in vaccinated individuals. Early, robust boosting of anti-spike protein antibodies was observed in vaccinated patients, however, these titers were significantly lower against B.1.617.2 as compared with the wildtype vaccine strain.

**Conclusion:** The mRNA vaccines are highly effective at preventing symptomatic and severe COVID-19 associated with B.1.617.2 infection. Vaccination is associated with faster decline in viral RNA load and a robust serological response. Vaccination remains a key strategy for control of COVID-19 pandemic.

## Background

Availability of effective vaccines against severe acute respiratory syndrome coronavirus 2 (SARS-CoV-2) within one year of the first report of coronavirus disease 2019 (COVID-19) is remarkable. Phase 3 clinical trials of messenger RNA (mRNA) vaccines have demonstrated 92-95% efficacy in preventing symptomatic infection and severe disease [1-4] and intensive vaccination programs have reduced infection and mortality rates in multiple settings [5-7].

Emerging variants of concern (VOCs), such as B.1.1.7 (Alpha in the World Health Organization classification), B.1.351 (Beta), P.1 (Gamma), and B.1.617.2 (Delta) exhibit varied sequence changes and alteration of amino acid sequences of the spike protein. This has led to concerns of viral immune evasion and decreased vaccine effectiveness. Furthermore, these VOCs have been shown to be more transmissible [8-10], and B.1.1.7 and B.1.617.2 has been associated with increased disease severity and hospitalization [11, 12]. B.1.617.2 has rapidly spread outside India, becoming the most frequently sequenced lineage worldwide by end of June 2021 [13]. Case series of vaccine-breakthrough infections have reported an over-representation by these VOCs [14, 15].

Understanding vaccine effectiveness in the context of VOCs requires granular data: which vaccines were administered, at what time point prior to infection, number of doses, and particularly which VOC has caused the infection. Important VOC-specific vaccination outcomes include severity of infection and vaccine effects on transmission.

The COVID-19 vaccination program was initiated in Singapore on 30 December 2020, with free vaccinations provided to all Singapore residents in phases, beginning with the elderly and those in high-risk occupations such as healthcare workers. Vaccines used are mRNA vaccines, Pfizer/BioNTech BNT162b2 and Moderna mRNA-1273. As of 19 July 2021, 6,837,200 vaccine doses had been administered and ∼2,792,430 individuals (47% of the total population) had completed the vaccination course [16]. In May 2021, B.1.617.2 became the dominant circulating variant based on local sequencing data. In this multi-center cohort study, we characterize the clinical features, virological and serological kinetics of patients with vaccine-breakthrough PCR-confirmed B.1.617.2 infection and compared them with unvaccinated patients.

## Methods

### Patient Recruitment

Adults aged ≥18 years with COVID-19 confirmed by positive SARS-CoV-2 PCR and admitted to any of the five study sites from 1 April to 14 June 2021 were screened. Patients with B.1.617.2 infection (identification methods delineated below) were included in this analysis. Vaccine-breakthrough infection was defined as PCR-confirmed COVID-19 with symptom onset or first positive PCR (whichever was earlier) ≥14 days following a second dose of BNT162b2 or mRNA-1273 vaccine. Incomplete vaccination was defined as receipt of one dose of these vaccines ≥14 days prior to symptom onset or first positive PCR. Patients who received non-mRNA vaccines or developed infection within 14 days after the first dose were excluded from this analysis. B.1.617.2 vaccine-breakthrough infections were compared with a retrospective cohort of unvaccinated patients with B.1.617.2 infection admitted to one study site.

### Data Collection

Clinical and laboratory data were collected from electronic medical records using a standardized data-collection form [17]. Laboratory data including cycle threshold (Ct) values from SARS-CoV-2 RT-PCR assays and serological results from Elecsys^®^ (Roche, Basel, Switzerland) Anti-SARS-CoV-2 chemiluminescent immunoassays [anti-nucleocapsid (anti-N) and anti-spike protein (anti-S)] and surrogate virologic neutralization test (sVNT) cPass™ (Genscript, NJ, USA) were recorded. cPass™ detects total neutralizing antibodies targeting the viral spike protein receptor-binding domain [18]. These tests were performed as part of routine clinical care.

### Additional Serologic testing

Serum samples from a subset of vaccine-breakthrough patients who had separately consented for specimen collection were additionally tested with a newly developed multiplex-sVNT assay using the Luminex platform. Further details can be found in the supplementary information.

### Viral RNA sequencing and VOC determination

SARS-CoV-2 PCR was performed using various commercially available assays in different clinical laboratories. As part of active genomic surveillance, whole genome sequencing (WGS) by National Public Health Laboratory is performed for all patients in Singapore with SARS-CoV-2 detected by RT-PCR with a Ct value less than 30. Pangolin COVID-19 Lineage Assigner and CoVsurver were used to assign lineage to each sequence. For individuals with PCR confirmed infection without available sequencing results, lineage was inferred based on epidemiological investigations by the Singapore Ministry of Health (MOH), and likely B.1.617.2 infections were included (i.e., clear epidemiologic link with patients with sequencing confirmed B.1.617.2 infection).

### Clinical Management

All individuals with confirmed COVID-19 (including asymptomatic cases) in Singapore are admitted to hospital for inpatient evaluation and isolation. Individuals with pneumonia requiring supplemental oxygen are treated with intravenous remdesivir, while dexamethasone and other agents were reserved for progressive infections per national guidelines [19]. Disease severity was stratified into asymptomatic, mild (no pneumonia on chest radiography), moderate (presence of pneumonia on chest radiography), severe (requiring supplemental oxygen), or critical (requiring intensive care unit [ICU] admission or mechanical ventilation). Collection of clinical data was censored on discharge from hospital.

### Statistical Analysis

For descriptive analysis, data were presented as median (interquartile range (IQR)) for continuous parameters and frequency (percentage) for categorical variables. Chi-square and Fisher’s exact tests were used to compared categorical variables, while for continuous variables, t-test was used for normal data and Mann-Whitney U test for non-normal data. For asymptomatic patients, the day of confirmatory COVID-19 diagnosis was denoted as day one of illness. For symptomatic patients, the day of symptom onset or the day of confirmatory COVID-19 diagnosis, whichever earlier, was denoted as day one of illness.

Previously reported risk factors for disease severity [20] were evaluated and included in a multivariate logistic regression model [21]. For serial Ct values, we fitted a generalized additive mixed model (GAMM) with a random intercept by patient. To investigate the effect of vaccination status on rate of increase of Ct value, we included fixed factors of vaccination status and day of illness with smoothing terms and interaction between these two fixed factors. We plotted Ct values with marginal effect of day of illness by vaccination status and 95% confidence intervals (CI) from the GAMM.

For analysis of cPass™ and anti-S titres we fitted a GAMM to serial titres with random intercept by patient in addition to fixed factor of day of illness with smoothing terms, separately for vaccine-breakthrough and unvaccinated patients infected with Delta variant. We plotted cPass™/anti-S titres with marginal effect of day of illness and 95%CI from GAMM for each group of vaccine-breakthrough and unvaccinated patients.

P-values less than 0.05 were considered statistically significant, and all tests were 2-tailed. Data analyses were performed using Stata Release 15 (StataCorp, College Station, TX) and R version 3.6.2 (R Foundation for Statistical Computing, Vienna, Austria).

### Ethical approval

Written informed consent was obtained from study participants of the multi-centre study approved by National Healthcare Group Domain Specific Review Board (NHG-DSRB) (Study Reference 2012/00917). Informed consent for retrospective data collection at National Centre for Infectious Diseases (NCID) was waived (NHG-DSRB reference number 2020/01122).

## Results

218 B.1.617.2 infections were identified across the five study sites (Supplementary Figure S1). Of these, 71 met the definition for vaccine-breakthrough. An additional 13 only received one dose ≥14 days prior to disease onset or received both doses but within 14 days of disease onset, while four had received a non-mRNA vaccine overseas. Majority of participants meeting study definition for vaccine-breakthrough had received two doses of BNT162b2 (n=66, 93%).

### Clinical Features

In line with Singapore’s national vaccination strategy wherein older adults were prioritized for vaccination, our vaccine-breakthrough cohort was of significantly older age; median age of 56 years (IQR:39-64) versus 39.5 (IQR:30-58) (p<0.001) (Table 1). Other baseline demographics were similar.

**Table 1:**
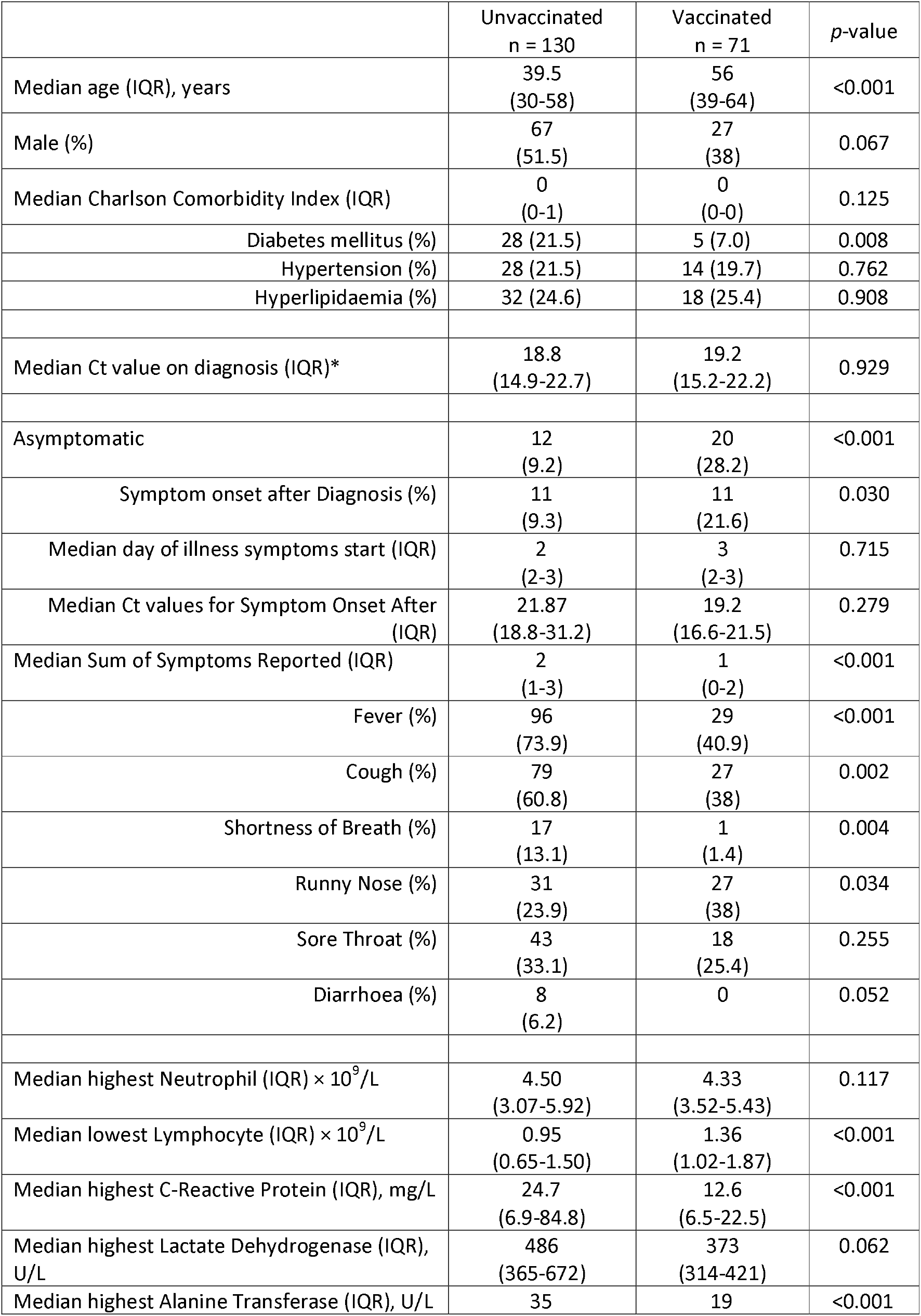
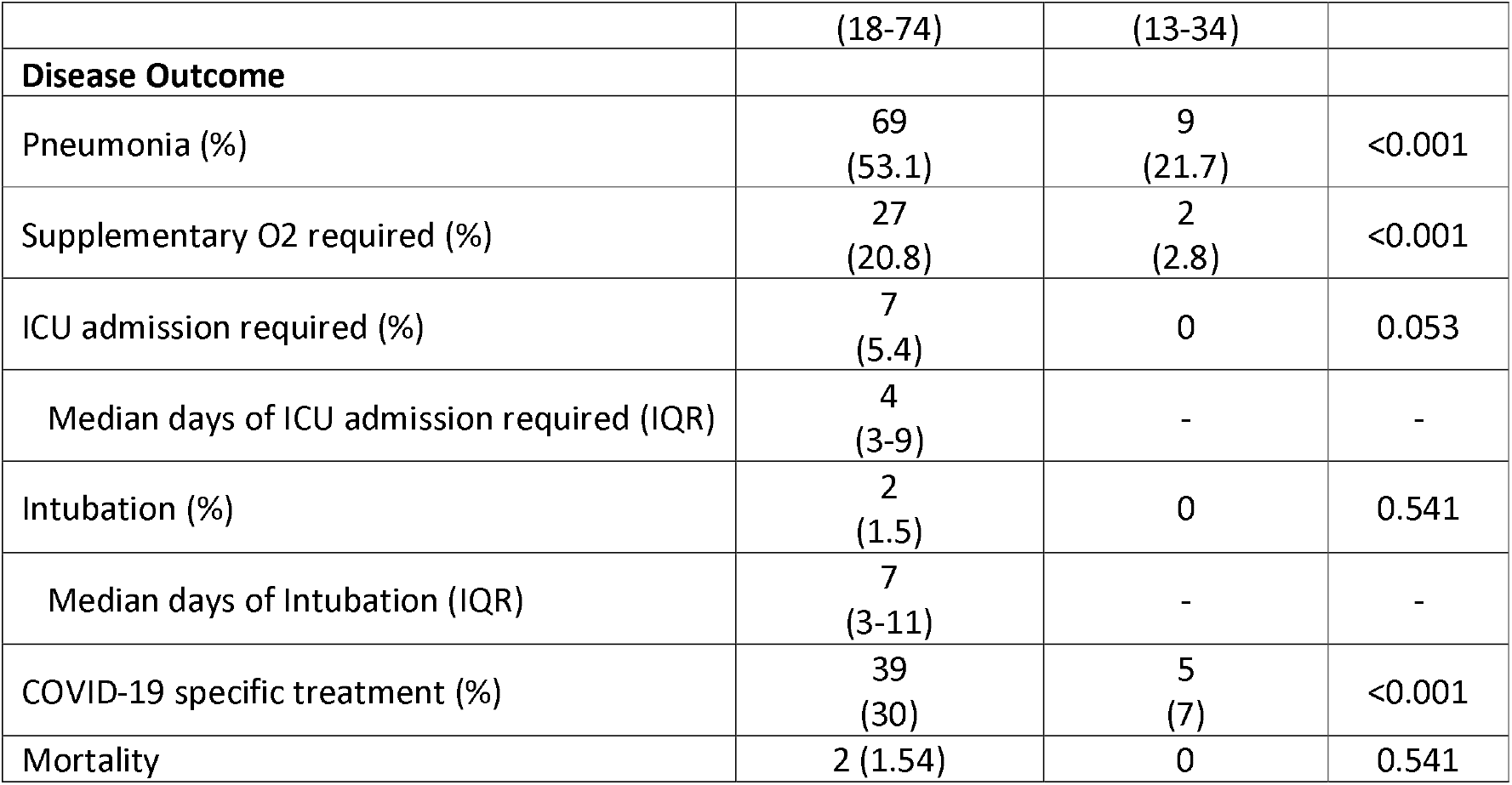
Baseline characteristics and disease outcome between unvaccinated and completed mRNA vaccination COVID-19 B1.617.2 infected patients

|  | Unvaccinated<br>n = 130 | Vaccinated<br>n = 71 | p-value |
| --- | --- | --- | --- |
| Median age (IQR), years | 39.5<br>(30-58) | 56<br>(39-64) | <0.001 |
| Male (%) | 67<br>(51.5) | 27<br>(38) | 0.067 |
| Median Charlson Comorbidity Index (IQR) | 0<br>(0-1) | 0<br>(0-0) | 0.125 |
| Diabetes mellitus (%) | 28 (21.5) | 5 (7.0) | 0.008 |
| Hypertension (%) | 28 (21.5) | 14 (19.7) | 0.762 |
| Hyperlipidaemia (%) | 32 (24.6) | 18 (25.4) | 0.908 |
| Median Ct value on diagnosis (IQR)* | 18.8<br>(14.9-22.7) | 19.2<br>(15.2-22.2) | 0.929 |
| Asymptomatic | 12<br>(9.2) | 20<br>(28.2) | <0.001 |
| Symptom onset after Diagnosis (%) | 11<br>(9.3) | 11<br>(21.6) | 0.030 |
| Median day of illness symptoms start (IQR) | 2<br>(2-3) | 3<br>(2-3) | 0.715 |
| Median Ct values for Symptom Onset After (IQR) | 21.87<br>(18.8-31.2) | 19.2<br>(16.6-21.5) | 0.279 |
| Median Sum of Symptoms Reported (IQR) | 2<br>(1-3) | 1<br>(0-2) | <0.001 |
| Fever (%) | 96<br>(73.9) | 29<br>(40.9) | <0.001 |
| Cough (%) | 79<br>(60.8) | 27<br>(38) | 0.002 |
| Shortness of Breath (%) | 17<br>(13.1) | 1<br>(1.4) | 0.004 |
| Runny Nose (%) | 31<br>(23.9) | 27<br>(38) | 0.034 |
| Sore Throat (%) | 43<br>(33.1) | 18<br>(25.4) | 0.255 |
| Diarrhoea (%) | 8<br>(6.2) | 0 | 0.052 |
| Median highest Neutrophil (IQR) $\times 10^9/L$ | 4.50<br>(3.07-5.92) | 4.33<br>(3.52-5.43) | 0.117 |
| Median lowest Lymphocyte (IQR) $\times 10^9/L$ | 0.95<br>(0.65-1.50) | 1.36<br>(1.02-1.87) | <0.001 |
| Median highest C-Reactive Protein (IQR), mg/L | 24.7<br>(6.9-84.8) | 12.6<br>(6.5-22.5) | <0.001 |
| Median highest Lactate Dehydrogenase (IQR), U/L | 486<br>(365-672) | 373<br>(314-421) | 0.062 |
| Median highest Alanine Transferase (IQR), U/L | 35 | 19 | <0.001 |

|  | (18-74) | (13-34) |  |
| --- | --- | --- | --- |
| **Disease Outcome** |  |  |  |
| Pneumonia (%) | 69<br>(53.1) | 9<br>(21.7) | <0.001 |
| Supplementary O2 required (%) | 27<br>(20.8) | 2<br>(2.8) | <0.001 |
| ICU admission required (%) | 7<br>(5.4) | 0 | 0.053 |
| Median days of ICU admission required (IQR) | 4<br>(3-9) | - | - |
| Intubation (%) | 2<br>(1.5) | 0 | 0.541 |
| Median days of Intubation (IQR) | 7<br>(3-11) | - | - |
| COVID-19 specific treatment (%) | 39<br>(30) | 5<br>(7) | <0.001 |
| Mortality | 2 (1.54) | 0 | 0.541 |

Vaccine-breakthrough patients were significantly more likely to be asymptomatic (28.2% versus 9.2%, p<0.001); and if symptomatic, had fewer number of symptoms (Table 1). Unvaccinated individuals had worse levels of known biomarkers associated with increased COVID-19 severity including lymphocyte count, C-reactive protein [CRP], lactate dehydrogenase [LDH] and alanine transferase [ALT]. Correspondingly, a higher proportion of the unvaccinated cohort had pneumonia, required supplementary oxygen and ICU admission compared with the vaccinated cohort. A broader analysis comparing unvaccinated versus those who had received at least one dose of vaccine (i.e. both vaccine-breakthrough and incomplete vaccination) demonstrated similar findings (Supplementary Table T1).

Multivariate logistic regression analysis for development of severe COVID-19 (defined by supplementary oxygen requirement) demonstrated that vaccination was protective with an adjusted odds ratio (aOR) of 0.073 (95% confidence interval [CI]):0.016-0.343) (p=0.001) (Table 2). Analysis comparing unvaccinated versus those who had received at least one dose of vaccine demonstrated similar findings (Supplementary Table T2). Multivariate logistic regression analysis for development of moderately severe COVID-19 (defined by development of pneumonia) also demonstrated that vaccination was protective with aOR of 0.069 (95%CI:0.027-0.180) (p<0.001) (Supplementary Table T3).

**Table 2:**
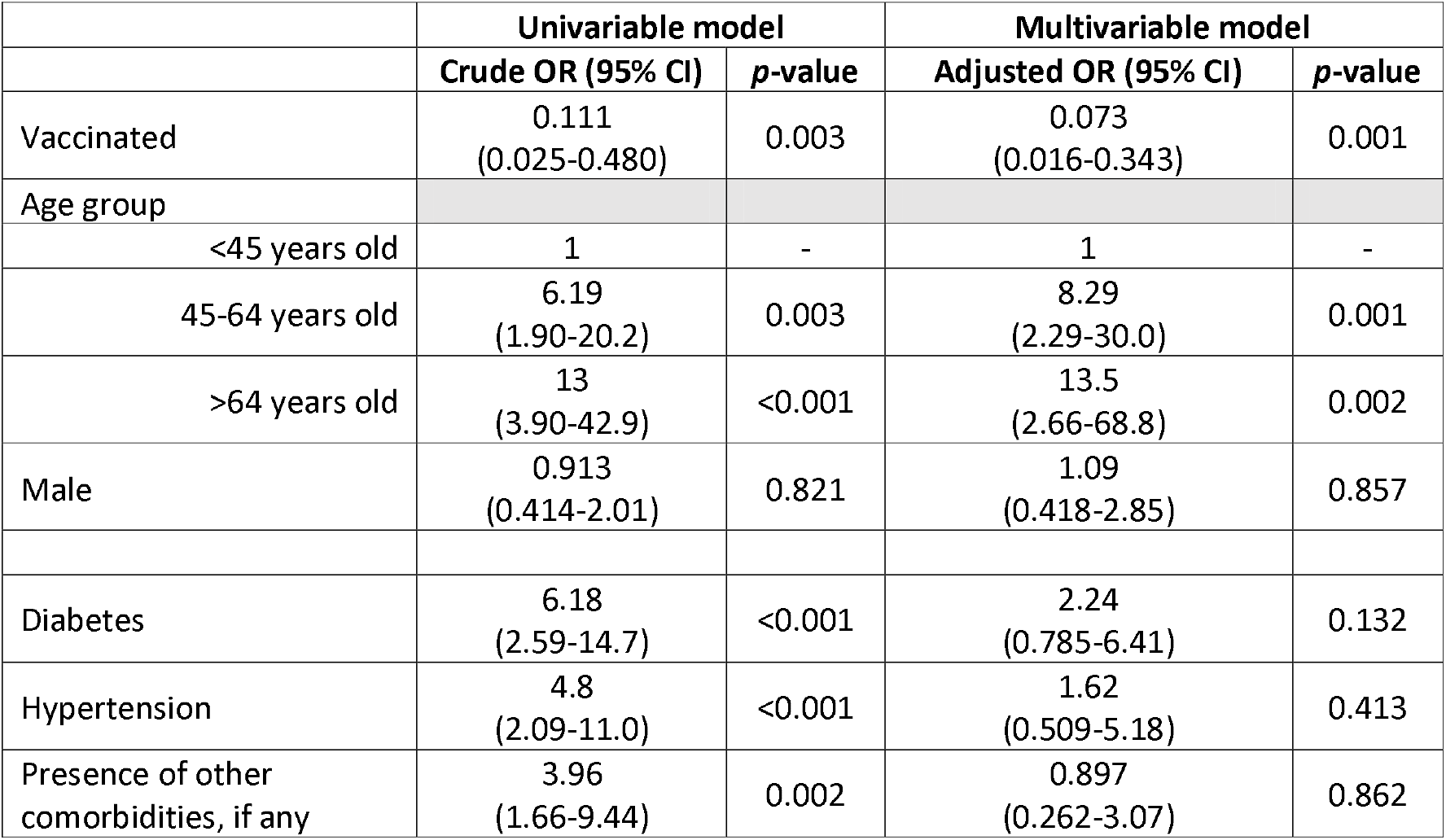
Odds ratio of candidate risk factors for development of severe COVID-19 for completed mRNA vaccination COVID-19 B1.617.2 infected patients. CI, confidence interval; OR, odds ratio

|  | Univariable model |  | Multivariable model |  |
| --- | --- | --- | --- | --- |
|  | Crude OR (95% CI) | *p*-value | Adjusted OR (95% CI) | *p*-value |
| Vaccinated | 0.111<br>(0.025-0.480) | 0.003 | 0.073<br>(0.016-0.343) | 0.001 |
| Age group |  |  |  |  |
| <45 years old | 1 | - | 1 | - |
| 45-64 years old | 6.19<br>(1.90-20.2) | 0.003 | 8.29<br>(2.29-30.0) | 0.001 |
| >64 years old | 13<br>(3.90-42.9) | <0.001 | 13.5<br>(2.66-68.8) | 0.002 |
| Male | 0.913<br>(0.414-2.01) | 0.821 | 1.09<br>(0.418-2.85) | 0.857 |
| Diabetes | 6.18<br>(2.59-14.7) | <0.001 | 2.24<br>(0.785-6.41) | 0.132 |
| Hypertension | 4.8<br>(2.09-11.0) | <0.001 | 1.62<br>(0.509-5.18) | 0.413 |
| Presence of other<br>comorbidities, if any | 3.96<br>(1.66-9.44) | 0.002 | 0.897<br>(0.262-3.07) | 0.862 |

### Virologic kinetics

Serial Ct values of individuals were analyzed as a surrogate marker for the viral load. The initial median initial Ct value did not differ between unvaccinated and fully vaccinated patients (unvaccinated median Ct 18.8 (14.9-22.7), vaccinated 19.2 (15.2-22.2), p=0.929). However, fully vaccinated patients had a faster rate of increase in Ct value over time compared with unvaccinated individuals, suggesting faster viral load decline (coefficient estimates for interaction terms ranged from 9.12 (standard error 3.75) to 12.06 (standard error 3.03); p-value <0.05 for each interaction terms) (Figure 1).

**Figure 1:**
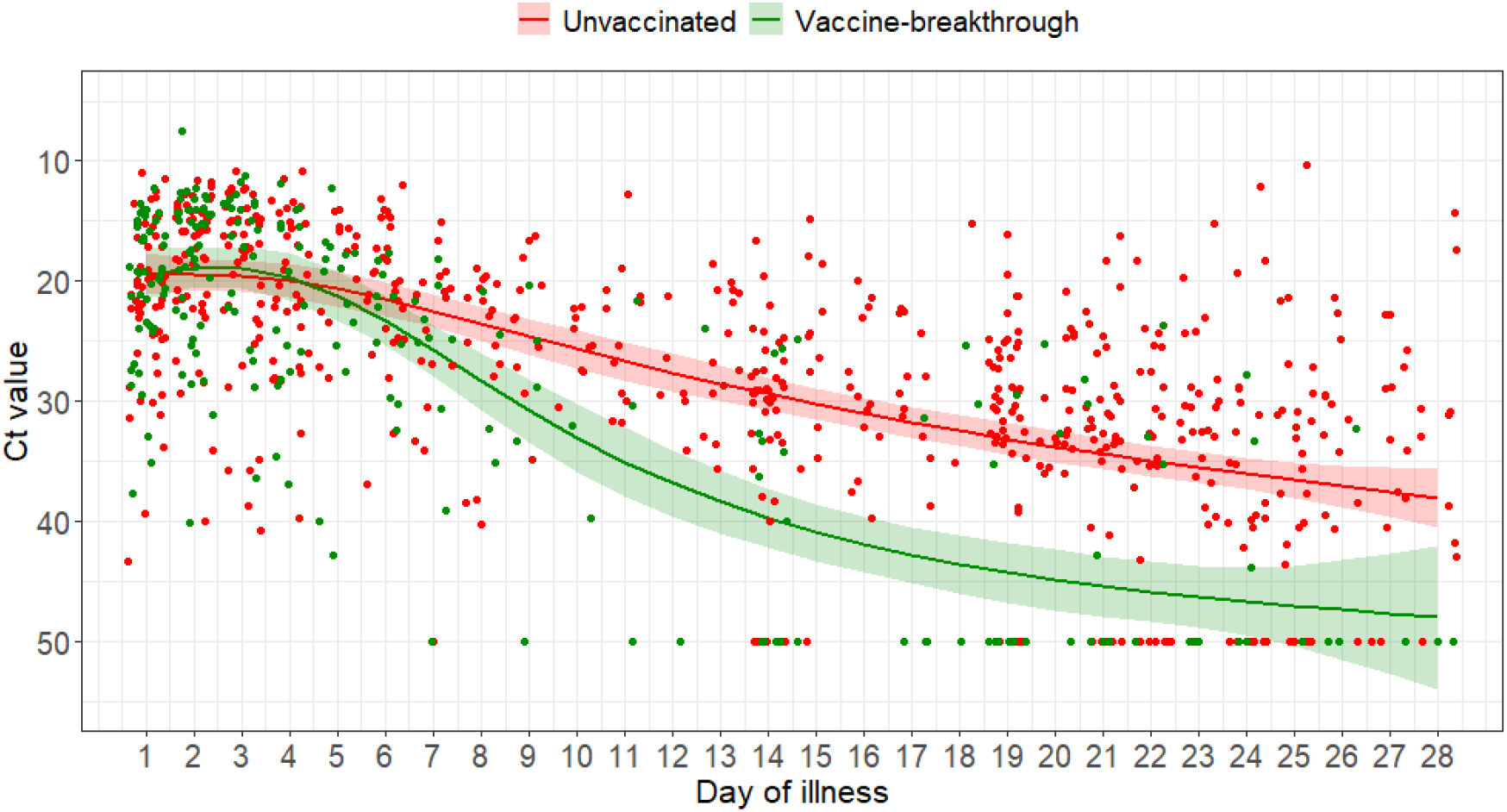
Scatterplot of Ct values and marginal effect of day of illness of COVID-19 B1.617.2 infected patients with 95% confidence intervals from generalized additive mixed model with interaction term between vaccination status and day of illness

### Serologic data

69 fully vaccinated individuals and 45 unvaccinated had serologic data available on record. 66/66 (100%) of vaccinated individuals had detectable S antibodies in week 1 of illness, while 7/45 (16%) of unvaccinated individuals did (Supplementary Figure S2). There was no difference in the proportion of individuals who seroconverted with the anti-N assay in week 1 (vaccinated 7/68 (10%) vs unvaccinated 11/107 (10%)) or week 2 (vaccinated 2/11 (18%), unvaccinated 4/20 (20%).

Analysis of sVNT with cPass indicated very high inhibition among vaccinated individuals in week 1 of illness (median 98.3% (IQR:91.0-99.4%)) which increased to 99.6% (IQR 99.3-99.9%) in week 2 (Figure 2A, 2B). Among unvaccinated individuals, median inhibition was below the 20% threshold at both week 1 and week 2. Among the 37 vaccinated individuals with a serum sample available for testing by the multiplex sVNT assay, titres were significantly higher against wildtype virus compared with B.1.617.2 and other VOCs (Figure 3). sVNT titres were lowest against B.1.617.2 and P.1 VOCs.

**Figure 2:**
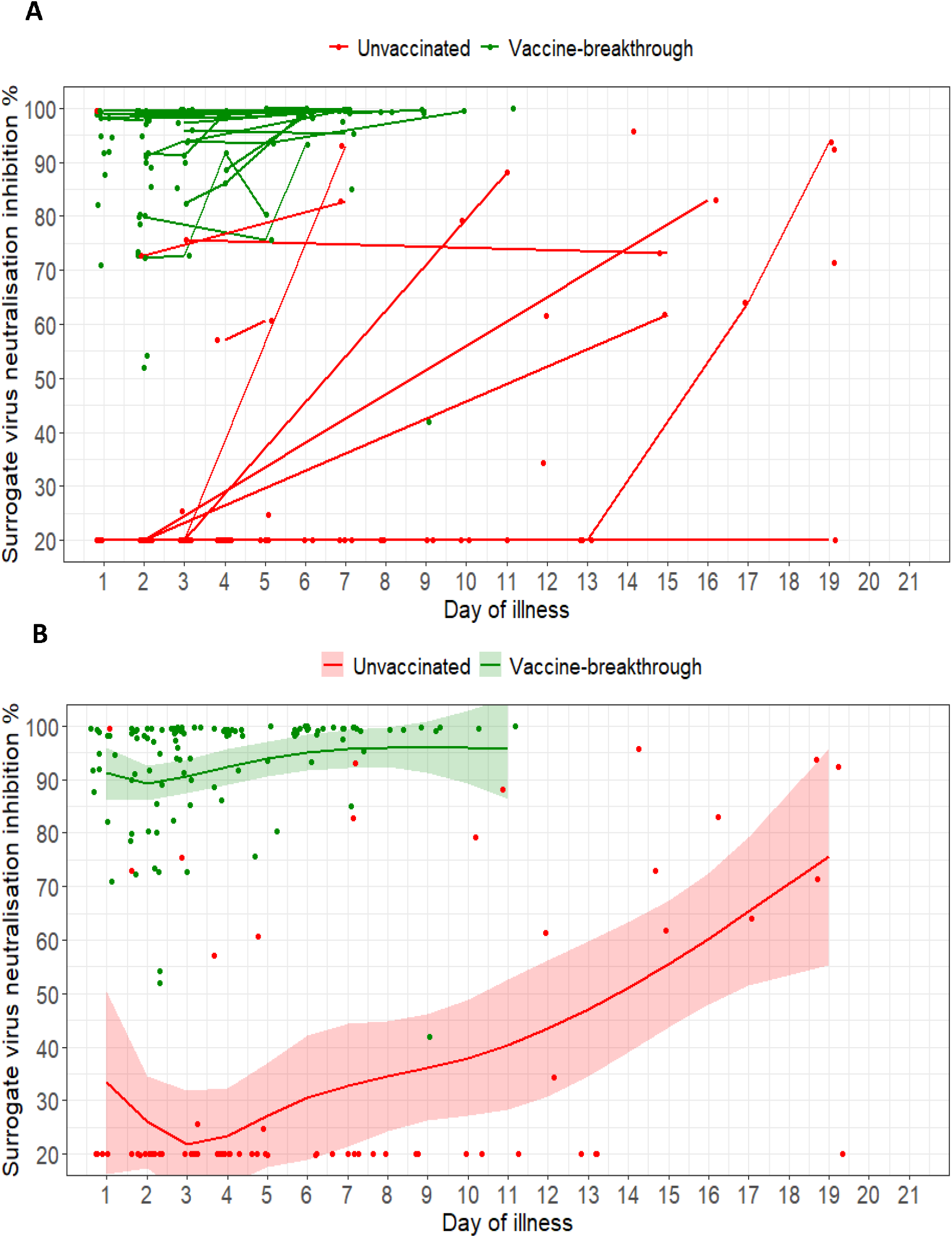
(A) Spaghetti plot of surrogate virus neutralisation (sVNT) inhibition % as measured by cPass; (B) Scatterplot of sVNT inhibition % and marginal effect of day of illness by vaccine-breakthrough and unvaccinated groups of COVID-19 B1.617.2 infected patients with 95% confidence intervals from generalized additive mixed models. For both plots, n=127; vaccine-breakthrough = 67, unvaccinated = 60

**Figure 3:**
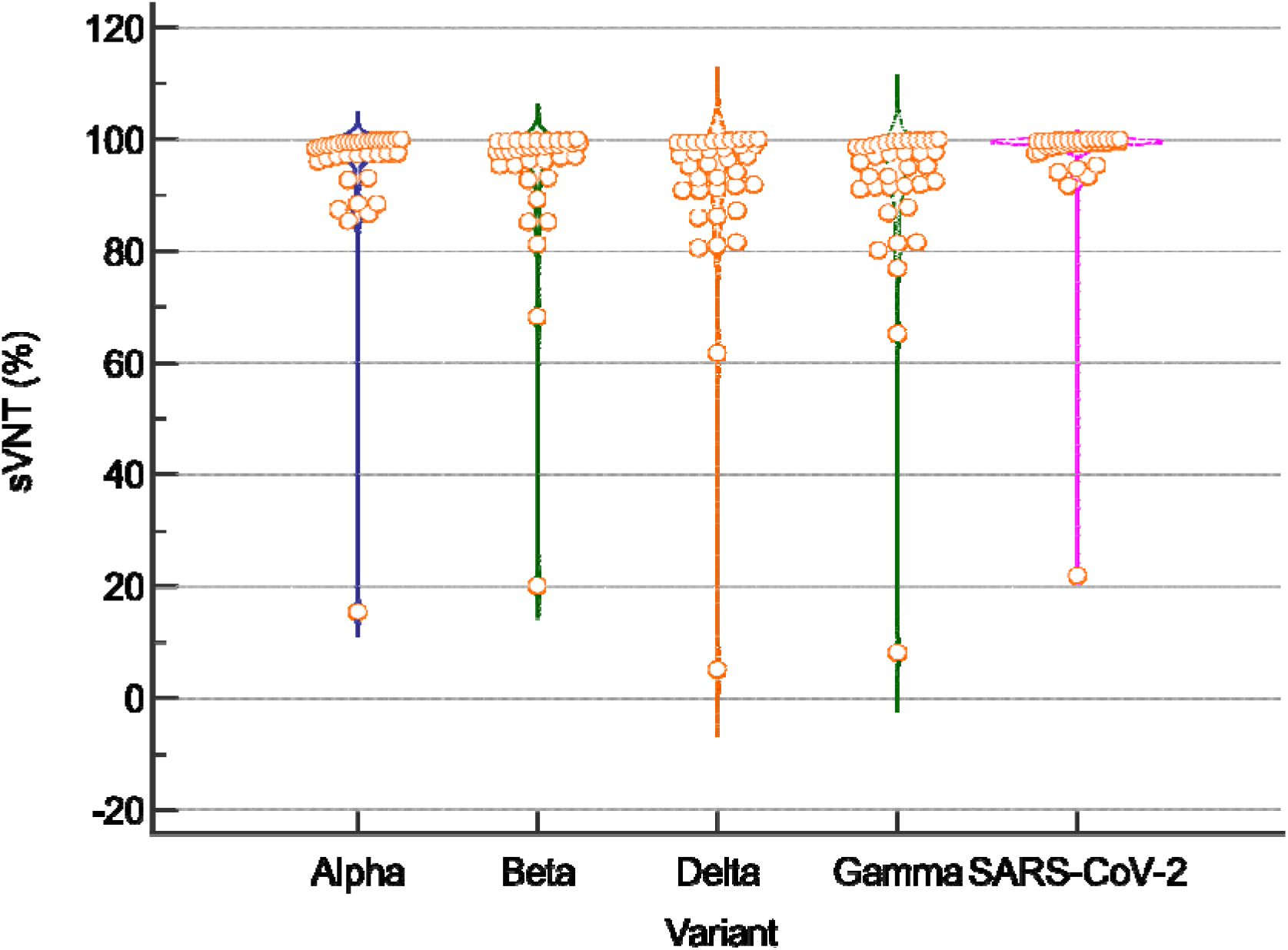
Violin plots of of surrogate virus neutralisation (sVNT) inhibition % against wildtype SARS-CoV-2 and the B.1.617.2 variant for 36 patients with vaccine-breakthrough infection (median day of sample collection from infection onset 6 days (inter-quartile range (IQR) 3-7). Titres against the four variants were significantly lower than against wildtype SARS-CoV-2 [median sVNT, B.1.1.7 98.5% (IQR: 96.3-99.5); B.1.351 98.2% (IQR: 95.3-99.5); B.1.617.2 96.0% (IQR: 90.9-99.3); P.1 95.5% (IQR: 91.3-98.9); Wildtype 99.4% (IQR: 98.5-99.7), Kruskal-Walis p-value = 0.00055, Post-hoc pairwise comparison (Conover) Wildtype versus each variant p<0.05]

## Discussion

In this study, we found that fully vaccinated patients had significantly lower odds of moderate or severe outcomes following infection by the SARS-CoV-2 VOC B.1.617.2. Vaccination was associated with lower peak measures of systemic inflammation, fewer symptoms, including more asymptomatic infection, and better clinical outcomes. Notably, in contrast to existing studies that showed lower viral load in vaccinated patients [22], initial viral load indicated by PCR Ct values was similar between vaccinated and unvaccinated patients with B.1.617.2. However, vaccinated patients appeared to clear viral load at a faster rate. Our serologic data suggest an early rapid rise in neutralizing and binding antibodies indicated by C-Pass and Roche anti-S antibodies, which may be evidence of memory immunity to COVID-19 vaccination on challenge with a breakthrough infection with B.1.617.2.

As part of active case finding and surveillance in Singapore, all patients with fever or respiratory symptoms, close contacts of confirmed cases, and newly arrived travelers are screened for COVID-19 using PCR. Additionally, high-risk individuals in frontline occupations or congregate settings are tested as part of routine surveillance. All confirmed COVID-19 cases are reported to MOH and admitted to a hospital for initial evaluation. As such, our hospitalized cohort uniquely captures the entire spectrum of disease severity of COVID-19 infection and provides granular data even for mild and asymptomatic vaccine-breakthrough infections, giving us the opportunity to analyze virologic and serologic kinetics of these patients.

The finding of diminished severity with B.1.617.2 infection in vaccinated individuals is reassuring and corroborates emerging data from the United Kingdom which have found that mRNA vaccination remains protective against symptomatic and severe disease[12, 23]. An observational cohort study conducted in Scotland suggested that ≥14 days after the second dose, BNT162b2 vaccine offered 92% vaccine effectiveness against presumptive non-B.1.617.2 infection and 79% protection against presumptive B.1.617.2 [24]. Protection associated with the ChAdOx1 nCoV-19 vaccine was 73% and 60% respectively. Although vaccine-breakthrough infections are increasingly reported, with the largest series to date in the United States reporting 10,262 breakthrough infections, a majority of these were mild (27% asymptomatic, 10% hospitalization, 2% mortality)[25]. Vaccine-breakthrough infections will continue to be observed, especially with genetic drift and selection pressures resulting in emergence of newer VOCs; however, it is likely that there will be a shift toward milder disease spectrum with more widespread implementation of vaccination programs.

To our knowledge, we provide the first data characterizing impact of vaccination on virologic kinetics by the B.1.617.2 variant. While initial Ct values were similar; the effect of vaccination with a more rapid decline in viral load (and hence shorter duration of viral shedding) has implications on transmissibility and infection control policy. A shorter duration of infectivity may allow a shorter duration of isolation for vaccinated individuals. Based on our data, it seems likely that vaccination reduces secondary transmission, though this needs to be further studied in larger community surveillance studies. Other studies found similar impact of vaccination on other variants. Pritchard and colleagues found that vaccinated individuals had higher Ct values compared with unvaccinated individuals in B.1.1.7 infections [7], while Levine-Tiefenbaum and colleagues similarly found a reduction in viral loads after BNT162b2 vaccine, though no data was provided on variant type [26].

There are several limitations to our study. Firstly, we only compared vaccine-breakthrough infections with unvaccinated COVID-19 patients. We did not study vaccinated individuals who had similar exposure risk but did not develop COVID-19 infection. We thus could not evaluate vaccine efficacy against asymptomatic infection. We also did not have detailed epidemiologic data to study the effect of vaccination on preventing secondary transmission.

Secondly, we could only obtain serologic tests after infection since patients were recruited after confirmation of infection. While active contact tracing and case finding in Singapore resulted in early identification of most COVID-19 cases, the first available serologic result was at a median of 2 (IQR:1-3) days of illness and antibody levels are likely to already have been boosted by natural infection. We thus could not evaluate the underlying immunologic mechanisms behind vaccine-breakthrough infection, e.g., diminished neutralizing antibody level or impaired cellular immunity. Further study should compare similarly exposed vaccinated individuals who develop breakthrough infection with those who do not, to elucidate the underlying drivers of susceptibility, which may enlighten us on how to optimize protection (e.g., through enhanced/boosted dosing schedules).

Thirdly, PCR testing was not standardized in a centralized laboratory, and instead conducted at each centre using different validated commercial assays. Ct values are only a surrogate measure of viral load and shedding. We did not evaluate viability of shed virus via viral culture. In addition, we only evaluated participants with mRNA vaccination, and thus our findings are restricted to mRNA vaccines and not all COVID-19 vaccines.

## Conclusion

mRNA vaccines against COVID-19 are protective against symptomatic infection and severe disease by the B.1.617.2 variant. Vaccinated individuals had a more rapid decline in viral load, which has implications on secondary transmission and public health policy. Rapid and widespread implementation of vaccination programs remains a key strategy for control of COVID-19 pandemic. Further studies should elucidate immunologic features driving vaccine-breakthrough infection to improve vaccine-induced protection.

## Data Availability

The datasets generated during and/or analysed during the current study are available from the corresponding author on reasonable request.

## Acknowledgments

We thank all clinical and nursing staff who provided care for the patients and staff in the Infectious Disease Research and Training Office of the National Centre for Infectious Diseases who assisted with data collection. We will also like to thank Jeremy Cutter at the National Public Health and Epidemiology Unit of National Centre for Infectious Diseases who assisted with data management on Ct values.

