## Supplementary material for "Virological and serological kinetics of SARS-CoV-2 Delta variant vaccine-breakthrough infections: a multi-center cohort study": S1, ST1, ST2, ST3, S2

**Supplementary Information**

**Additional Serologic testing**

Briefly, AviTag-biotinylated receptor binding domain (RBD) proteins from wildtype SARS-CoV-2 and four VOCs (Alpha, Beta, Delta, Gamma) were coated on a MagPlex Avidin microsphere (Luminex) at 5 µg/1 million beads. RBD-coated microspheres (600 beads/antigen) were pre-incubated with serum at a final 1:20 or greater for 1h at 37°C with 800 rpm agitation. After 1 h incubation, 50 µl of phycoerythrin (PE)-conjugated hACE2 (GenScript, 1 µg/ml) were added to the well and incubated for 30 min at 37°C with agitation, followed by two PBS-1% BSA washes. The final readings were acquired using the MAGPIX system.

NCID, n = 130

218 participants

n=76

Unvaccinated

n = 130

Vaccinated

n = 88

NCID, n = 76

SGH, n = 5

NUH n = 3

CGH, n = 2

SKH, n = 2

Supplementary Figure S1: Study flowchart of participant recruitment from each study site: National Centre for Infectious Diseases (NCID), Singapore General Hospital (SGH); National University Hospital (NUH), Changi General Hospital (CGH) and Sengkang Hospital (SKH). Of the 88 vaccinated individuals: 71 had received 2 doses of a mRNA vaccine, 13 had received one dose of a mRNA vaccine and 4 received a non-mRNA vaccine.

|  | Unvaccinated  n = 130 | Vaccinated  n = 84 | *p*-value |
| --- | --- | --- | --- |
| **Baseline Characteristics** |  |  |  |
| Median age (IQR), years | 39.5  (30-58) | 57  (40.5-64) | <0.001 |
| Male (%) | 67  (51.5) | 33  (39.3) | 0.079 |
| Median Charlson Comorbidity Index (IQR) | 0  (0-1) | 0  (0-0) | 0.073 |
| Diabetes mellitus (%) | 28 (21.5) | 9 (10.7) | 0.041 |
| Hypertension (%) | 28 (21.5) | 18 (21.4) | 0.985 |
| Hyperlipidaemia (%) | 32 (24.6) | 25 (29.8) | 0.406 |
| Median Ct value on diagnosis (IQR) ꭞ | 18.8  (14.9-22.7) | 19.0  (15.2-22.9) | 0.960 |
| Asymptomatic | 12  (9.2) | 21  (25) | 0.002 |
| Symptom onset after Diagnosis (%) | 11  (9.3) | 13  (20.6) | 0.033 |
| Median day of illness symptoms start (IQR) | 2  (2-3) | 2  (2-3) | 1.000 |
| Median Ct values for Symptom Onset After (IQR) | 21.87  (18.8-31.2) | 19.3  (16.6-22.2) | 0.356 |
| Median number of Symptoms Reported (IQR) | 2  (1-3) | 1  (0-2.5) | <0.001 |
| Fever (%) | 96  (73.9) | 39  (46.4) | <0.001 |
| Cough (%) | 79  (60.8) | 30  (35.7) | <0.001 |
| Shortness of Breath (%) | 17  (13.1) | 3  (3.6) | 0.028 |
| Runny Nose (%) | 31  (23.9) | 30  (35.7) | 0.060 |
| Sore Throat (%) | 43  (33.1) | 22  (26.2) | 0.285 |
| Diarrhoea (%) | 8  (6.2) | 1  (1.2) | 0.093 |
| Median highest Neutrophil (IQR) × 10^9^/L | 4.50  (3.07-5.92) | 4.31  (3.58-5.44) | 0.881 |
| Median lowest Lymphocyte (IQR) × 10^9^/L | 0.95  (0.65-1.50) | 1.41  (1-1.85) | <0.001 |
| Median highest C-Reactive Protein (IQR), mg/L | 24.7  (6.9-84.8) | 12.9  (6.1-22.7) | 0.001 |
| Median highest Lactate Dehydrogenase (IQR), U/L | 486  (365-672) | 377.5  (317-432) | <0.001 |
| Median highest Alanine Transferase (IQR), U/L | 35  (18-74) | 19  (13-34) | <0.001 |
| **Disease Outcome** | | | |
| Pneumonia (%) | 69  (53.1) | 16  (19.0) | <0.001 |
| Supplementary O_2_ required (%) | 27  (20.8) | 4  (4.8) | 0.001 |
| ICU admission required (%) | 7  (5.4) | 0 | 0.044 |
| Median days of ICU admission required (IQR) | 4  (3-9) | - | - |
| Intubation (%) | 2  (1.5) | 0 | 0.521 |
| Median days of Intubation (IQR) | 7  (3-11) | - | - |
| COVID-19 specific treatment (%) | 39  (30) | 9  (10.7) | 0.001 |
| Mortality | 2 (1.54) | 0 | 0.521 |

ꭞ Data available only for 122 unvaccinated cases and 77 vaccinated cases

**Supplementary Table ST1:** Baseline characteristics and disease outcomes of unvaccinated versus vaccinated COVID-19 B1.617.2 infected patients who received at least one dose of a mRNA vaccine 14 days prior. Four individuals who received a non-mRNA vaccine were not included.

|  | **Univariable model** | | **Multivariable model** | |
| --- | --- | --- | --- | --- |
|  | **Crude OR (95% CI)** | ***p*-value** | **Adjusted OR (95% CI)** | ***p*-value** |
| Vaccinated | 0.191  (0.064-0.567) | 0.003 | 0.116  (0.036-0.373) | <0.001 |
| Age group |  |  |  |  |
| <45 years old | 1 | - | 1 | - |
| 45-64 years old | 5.91  (1.81-19.0) | 0.003 | 8.15  (2.30-28.9) | 0.001 |
| >64 years old | 12.5  (3.81-41.0) | <0.001 | 10.6  (2.21-50.6) | 0.003 |
| Male | 0.929  (0.432-2.00) | 0.850 | 1.04  (0.415-2.59) | 0.940 |
| Diabetes | 5.73  (2.49-13.2) | <0.001 | 2.16  (0.798-5.84) | 0.130 |
| Hypertension | 4.60  (2.06-10.3) | <0.001 | 1.64  (0.543-4.94) | 0.381 |
| Presence of other comorbidities, if any | 4.36  (1.91-9.95) | <0.001 | 1.38  (0.444-4.31) | 0.576 |

**Supplementary Table ST2:** Odds ratio of candidate risk factors of development of severe COVID-19 for vaccinated COVID-19 B1.617.2 infected patients who received at least one dose of a mRNA vaccine 14 days prior. CI, confidence interval; OR, odds ratio

|  | **Univariable model** | | **Multivariable model** | |
| --- | --- | --- | --- | --- |
|  | **Crude OR (95% CI)** | ***p*-value** | **Adjusted OR (95% CI)** | ***p*-value** |
| Vaccinated | 0.128  (0.059-0.280) | <0.001 | 0.069  (0.027-0.180) | <0.001 |
| Age group |  |  |  |  |
| <45 years old | 1 |  | 1 |  |
| 45-64 years old | 2.63  (1.35-5.15) | 0.005 | 5.59  (2.21-14.2) | <0.001 |
| >64 years old | 3.87  (1.78-8.44) | 0.001 | 5.91  (1.66-21.1) | 0.006 |
| Male | 1.14  (0.644-2.01) | 0.659 | 1.05  (0.516-2.14) | 0.892 |
| Diabetes | 6.78  (2.87-16.0) | <0.001 | 3.08  (1.04-9.08) | 0.041 |
| Hypertension | 2.59  (1.30-5.19) | 0.007 | 0.888  (0.303-2.59) | 0.828 |
| Presence of other comorbidities, if any | 4.26  (1.94-9.36) | <0.001 | 1.75  (0.606-5.05) | 0.301 |

**Supplementary Table ST3:** Odds ratio of candidate risk factors of development of moderately severe COVID-19 for completed mRNA vaccination COVID-19 B1.617.2 infected patients. CI, confidence interval; OR, odds ratio

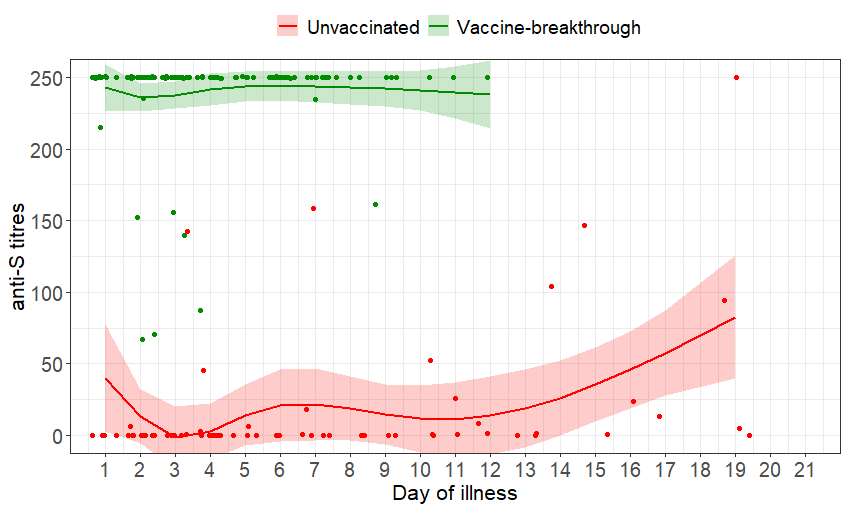

**Supplementary Figure S2:** Scatterplot of anti-S titres and marginal effect of day of illness by vaccine-breakthrough and unvaccinated groups of COVID-19 B1.617.2 infected patients with 95% confidence intervals from generalized additive mixed models. n=128; vaccine-breakthrough = 698, unvaccinated = 59
